# Assessing the genetic contribution of cumulative behavioral factors associated with longitudinal type 2 diabetes risk highlights adiposity and the brain-metabolic axis

**DOI:** 10.1101/2024.01.30.24302019

**Authors:** Nuno R. G. Carvalho, Yixuan He, Patrick Smadbeck, Jason Flannick, Josep M. Mercader, Miriam Udler, Arjun K Manrai, Jordi Moreno, Chirag J. Patel

## Abstract

While genetic factors, behavior, and environmental exposures form a complex web of interrelated associations in type 2 diabetes (T2D), their interaction is poorly understood. Here, using data from ∼500K participants of the UK Biobank, we identify the genetic determinants of a “polyexposure risk score” (PXS) a new risk factor that consists of an accumulation of 25 associated individual-level behaviors and environmental risk factors that predict longitudinal T2D incidence. PXS-T2D had a non-zero heritability (h^2^ = 0.18) extensive shared genetic architecture with established clinical and biological determinants of T2D, most prominently with body mass index (genetic correlation [r_g_] = 0.57) and Homeostatic Model Assessment for Insulin Resistance (r_g_ = 0.51). Genetic loci associated with PXS-T2D were enriched for expression in the brain. Biobank scale data with genetic information illuminates how complex and cumulative exposures and behaviors as a whole impact T2D risk but whose biology have been elusive in genome-wide studies of T2D.

---

Diabetes is a complex disease with many genetic, environmental, and behavioral factors hypothesized to have a role in etiology and disease progression. However, most studies focus on a single modality and the complex interplay between these factors across domains in diabetes onset has not been systematically characterized.^1–3^ While genetic and environmental factors are traditionally modeled as independent axes of physiological variation, they are, in fact, correlated.^1,2^

Work to uncover the correlation between genetic, environment, and behavioral factors has highlighted the importance of studying the “exposome.”^4^ The exposome is a classification of non-genetically measured variables belonging to *domains* of lifestyle behavior (e.g., as diet, physical activity, and smoking), social standing (e.g., income, education), and physical-chemical (e.g., pollution)^5^.^6–8^ Previously, we investigated domains of the exposome such as lifestyle behaviors and social factors, in prediction of incident type 2 diabetes (T2D).^9,10^ We called individual factors of the exposome within the domains “*exposures.*” We developed a “polyexposure score” (PXS) composed from 12 individual-level exposures that were indicators of lifestyle, social standing, and physical-chemical factors, executing a longitudinal exposure-wide association study (ExWAS) to identify variables with robust statistical support.^6^ The PXS played a complementary and independent role to a polygenic risk score (PRS) and traditional clinical risk factors (e.g., glycated hemoglobin A1C), body mass index [BMI], blood pressure, family history of disease, and cholesterol levels) in predicting incident T2D. Finding the role of antecedent exposures in diabetes physiology would complement recent advances in elucidating the biological function of genetic findings from large-scale GWAS.^11,12^ However, the mechanistic role of the PXS in T2D and its genetic architecture have not been interrogated.

In the following, we derive a new cumulative risk factor for T2D, called the PXS-T2D, and investigate the extent to which the composite PXS for incident T2D (PXS-T2D) and its individual-level behaviors are genetically heritable. We also investigate whether their genetic architectures are similar to that of clinical risk factors for disease. We focus on the subset of the exposome pertaining to individually-measured behaviors and study the phenomena of gene-exposure correlation (e.g. ^13^ ) to understand the relationship between the totality of exposure behaviors, as ascertained by the PXS, in T2D. We estimate the heritability and genetic correlations between behaviors and metabolic measurements and map associated variants to genes and tissue expression patterns, revealing the PXS is heritable and shares genetic architecture with antecedents of the disease and its complications. We show that elucidation of gene-exposure correlation at biobank scale enhances the identification of causal connections between exposures and physiological factors that are elusive in current day diabetes research.

## Results

Here, we systematically investigated the network of type 2 diabetes etiology (Figure 1A-C). We expanded on the model of He et al.^9^ (Figure 1B) by estimating the longitudinal risk for diabetes as a function of multiple behavior factors, creating a new cumulative score called PXS-T2D (Figure 1A, C). Next, we calculated the correlation between PXS-T2D and the polygenic risk score as well as the genetic correlation between PXS-T2D and clinical risk factors of type 2 diabetes (Figure 1B). We identified the genetic variants associated with the PXS-T2D and its component behaviors, for which we also compute genetic correlations with clinical risk factors (Figure 1C). The mean time to T2D diagnosis (incident T2D) was 2,741 days (7.51 years, range: 2–5242 days, standard deviation: 1174.4 days).

**Fig. 1.**
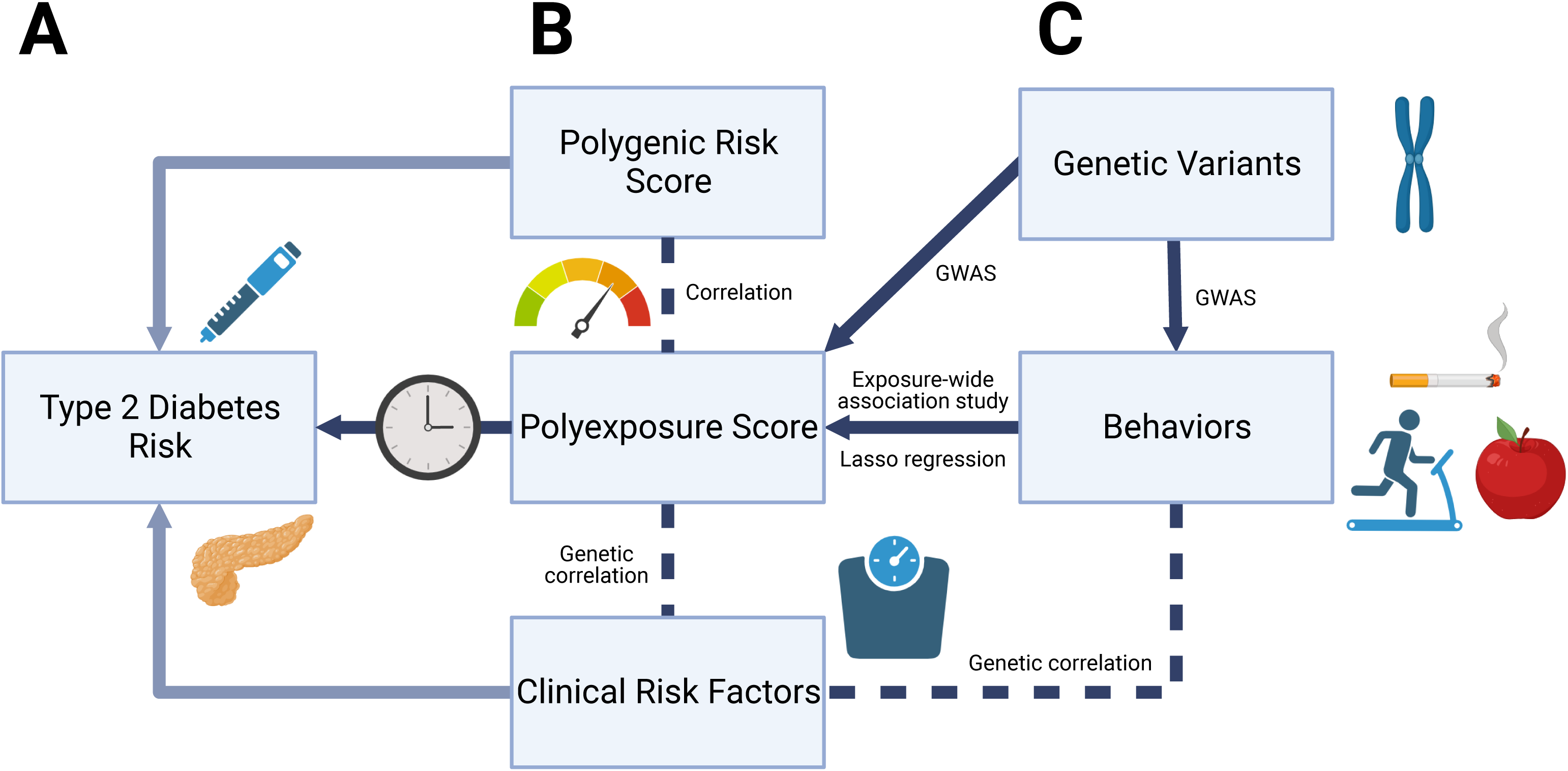
Model for the etiology of type 2 diabetes. Our model for type 2 diabetes (T2D) risk (A). The central columns (B) consists of the polygenic risk score (PRS), polyexposure score (PXS-T2D), and clinical risk factors (CRFs), which were considered in He et al.^9^ to predict incident T2D in the UK Biobank participants. In this paper, we investigate the genetic components of PXS-T2D and the 25 behavioral components that make it up (C), and how they correlate with clinical risk factors. Arrows shown in a darker shade of blue consist of relationships explored in this paper, and the methods employed are listed.

### Exposure-wide association study and PXS derivation

After false-discovery rate (FDR) correction,^14^ 84 variables remained with FDR-adjusted p-value less than 0.05 (Table S1). The final PXS-T2D model consisted of linear weights for 25 behavior variables in addition to age, sex, and 40 genetic principal components (Table S2) and predicted future diabetes with an area under the curve (AUC) of 0.74. Factors in the PXS-T2D included alcohol intake (Hazard Ratio = 1.2, p = 2 x 10^-43^), frequency of daytime naps (HR = 1.25, p = 6 x 10^-15^), current smoker (vs. non-smokers; HR = 1.2, p = 1 x 10^-10^), diet variety (HR = 1.3, p = 7 x 10^-15^) and time watching television (HR = 1.2, p = 3 x 10^-8^) associated with increased risk of future T2D. We also found factors such as walking pace (HR = 0.7, p = 4 x 10^-34^) and moderate physical activity (HR = 0.97, p = 9 x 10^-5^) associated with decreased risk for T2D. We note these were independently associated with T2D. The 25 behavioral exposures were weakly correlated with each other (Figure S1).

### The Polyexposure Score (PXS) is weakly correlated with the T2D Polygenic Risk Score, but is causally associated with Type 2 Diabetes

We used the PRS for T2D present in the UK Biobank^15^ to compute the correlation between the PRS and PXS for T2D. The correlation between the two variables was low, but significant given the large sample size (r = 0.0422, p < 2.2 x 10^-16^, Figure S2).

We were next interested in the potential causal relationship between the PXS-T2D and T2D. We used an approach called “Mendelian Randomization,”^16^ a method that associates genetic variants as proxies for exposures to disease outcomes, mitigating confounding that is common in observational/non-experimental studies. We input GWAS-significant SNP summary statistics for PXS-T2D (obtained via a GWAS in the UK Biobank, results discussed below) and GWAS summary statistics for T2D incidence in an inverse variance weighted Mendelian Randomization.^17^ PXS-T2D was associated causally for incident T2D (effect size = 0.072, 95% CI [0.065, 0.079], p < 1E-3). We also tested the reverse direction and had no statistically significant evidence to support that the association was reverse causal (effect size = -0.176 [-0.389, 0.037], p = 0.104). When using a broader definition for T2D that included cases before the first assessment, we found T2D to have a slightly stronger and significant causal association with PXS-T2D (effect size = 0.082 [0.030, 0.133], p = 0.002).

### PXS-T2D and its individual component exposure behaviors are heritable

We estimated SNP-based heritabilities for PXS-T2D and its 25 behavioral components. PXS-T2D has a heritability of 0.184 (95% CI [0.178, 0.190]), greater than the heritability of broadly defined (both incident and cross-sectional cases combined) T2D (h^2^=0.109, [0.106, 0.113]) and of incident T2D (h^2^=0.038, [0.034, 0.041]). We estimated the heritability of BMI-adjusted PXS-T2D and found a modest decrease in heritability compared to its non-adjusted counterpart (h^2^ = 0.144 [0.138, 0.149] vs. h^2^ = 0.184).

The heritability of 25 individual behaviors ranged from h^2^ = 0.0127 [0.00946, 0.0159] for “Renting for private landlord or letting agency” to h^2^ = 0.143 [0.138, 0.147] for “Time spent watching television.” The most heritable behaviors also included “Alcohol intake frequency” (h^2^ = 0.120, [0.116, 0.124]) and “Nap during day” (h^2^ = 0.119, [0.115, 0.123]). Heritability of all behaviors are in Table 1. No individual behavior had a higher heritability than PXS-T2D, even after adjusting it for BMI.

**Table 1:** Number of samples collected per time interval.

| Behavior | h<sup>2</sup> | h<sup>2</sup> 95% C.I. | Lambda | Significant SNPs | Significant Genes | Significant Loci |
| --- | --- | --- | --- | --- | --- | --- |
| **PXS for T2D onset** | 0.185 | [0.176, 0.195] | 1.223 | 6306 | 342 | 88 |
| Time watching TV | 0.143 | [0.136, 0.151] | 1.238 | 7054 | 243 | 81 |
| Alcohol intake | 0.122 | [0.115, 0.129] | 1.208 | 6868 | 316 | 65 |
| Daytime Naps | 0.119 | [0.112, 0.126] | 1.200 | 12325 | 184 | 91 |
| Walking pace | 0.105 | [0.098, 0.112] | 1.181 | 2722 | 166 | 39 |
| Sleep duration | 0.102 | [0.095, 0.108] | 1.185 | 8778 | 156 | 59 |
| Water intake | 0.094 | [0.087, 0.101] | 1.147 | 1549 | 107 | 28 |
| Dried fruit intake | 0.094 | [0.086, 0.101] | 1.157 | 2652 | 97 | 33 |
| Current smoker | 0.089 | [0.082, 0.096] | 1.155 | 3038 | 74 | 32 |
| White bread consumption | 0.089 | [0.082, 0.096] | 1.155 | 1756 | 71 | 24 |
| Time using phone | 0.087 | [0.080, 0.093] | 1.145 | 1699 | 43 | 29 |
| Tea intake | 0.087 | [0.080, 0.093] | 1.143 | 5269 | 153 | 28 |
| Previous smoker | 0.084 | [0.077, 0.090] | 1.146 | 2876 | 34 | 30 |
| Bread intake | 0.077 | [0.070, 0.084] | 1.139 | 2381 | 67 | 30 |
| Moderate physical activity frequency | 0.064 | [0.058, 0.071] | 1.113 | 1766 | 85 | 12 |
| Diet variety | 0.064 | [0.058, 0.070] | 1.122 | 977 | 48 | 12 |
| Vigorous physical activity frequency | 0.061 | [0.054, 0.067] | 1.102 | 1444 | 15 | 11 |
| Rent social housing | 0.060 | [0.054, 0.067] | 1.129 | 297 | 8 | 12 |
| Never eat sugar | 0.056 | [0.050, 0.062] | 1.100 | 26 | 21 | 5 |
| Stair climbing frequency | 0.055 | [0.049, 0.061] | 1.101 | 227 | 33 | 9 |
| No recent dietary change | 0.053 | [0.046, 0.059] | 1.102 | 383 | 28 | 8 |
| Poultry intake | 0.049 | [0.042, 0.055] | 1.100 | 449 | 4 | 3 |
| No disability benefits | 0.039 | [0.033, 0.045] | 1.079 | 92 | 9 | 9 |
| Illness-induced dietary change | 0.030 | [0.024, 0.036] | 1.062 | 18 | 1 | 2 |
| Brown bread consumption | 0.017 | [0.011, 0.022] | 1.036 | 14 | 2 | 2 |
| Rent private housing | 0.013 | [0.007, 0.018] | 1.054 | 287 | 13 | 22 |

### Genetic markers associated with PXS-T2D and behaviors

Using the PXS-T2D model, we computed the PXS score for each subject in the UK Biobank and performed a Genome-Wide Association Study (GWAS) for PXS-T2D and all its component exposure behaviors (Table 1). For PXS-T2D, we identified 6,306 SNPs below the genome-wide significance threshold (p < 5 x 10^-8^), which mapped to 342 genes and 88 genomic loci (Figure 2, Table S3). The most significant locus was rs13135092 (beta = -0.0276, p = 3.5 x 10^-20^), an intronic variant found in chromosome 4. This SNP maps closely onto the *SLC39A8* gene, which encodes the manganese solute carrier ZIP8 and is associated with neurological disorders like congenital disorder of glycosylation type II.^18^ *SLC39A8* was not found to be associated with T2D in the T2DKP^19,20^ (z = 1.53, p = 0.063), but the gene includes SNPs that have achieved GWAS-significant association with body mass index (BMI), diastolic blood pressure, triglycerides, and high density lipoprotein (HDL) cholesterol, and Insulin Growth Factor-1 (IGF-1).^19^ *SLC39A8* was also associated with behavior variables such as walking pace (beta = 0.0205, p = 9.2 x 10^-16^), Alcohol intake (beta = -0.0384, p = 7.8 x 10^-11^), “Sleep duration” (beta = 0.0297, p = 1.5 x 10^-10^), and “Time watching TV” (beta = -0.0192, p = 1.9 x 10^-9^); all associations were directionally consistent with those for PXS-T2D and T2D. Another gene within the same locus as rs13135092 is the *BANK1* gene, associated with systemic sclerosis^21^ and other autoimmune diseases^22^ but not T2D in the T2DKP (p = 0.823).

**Fig. 2.**
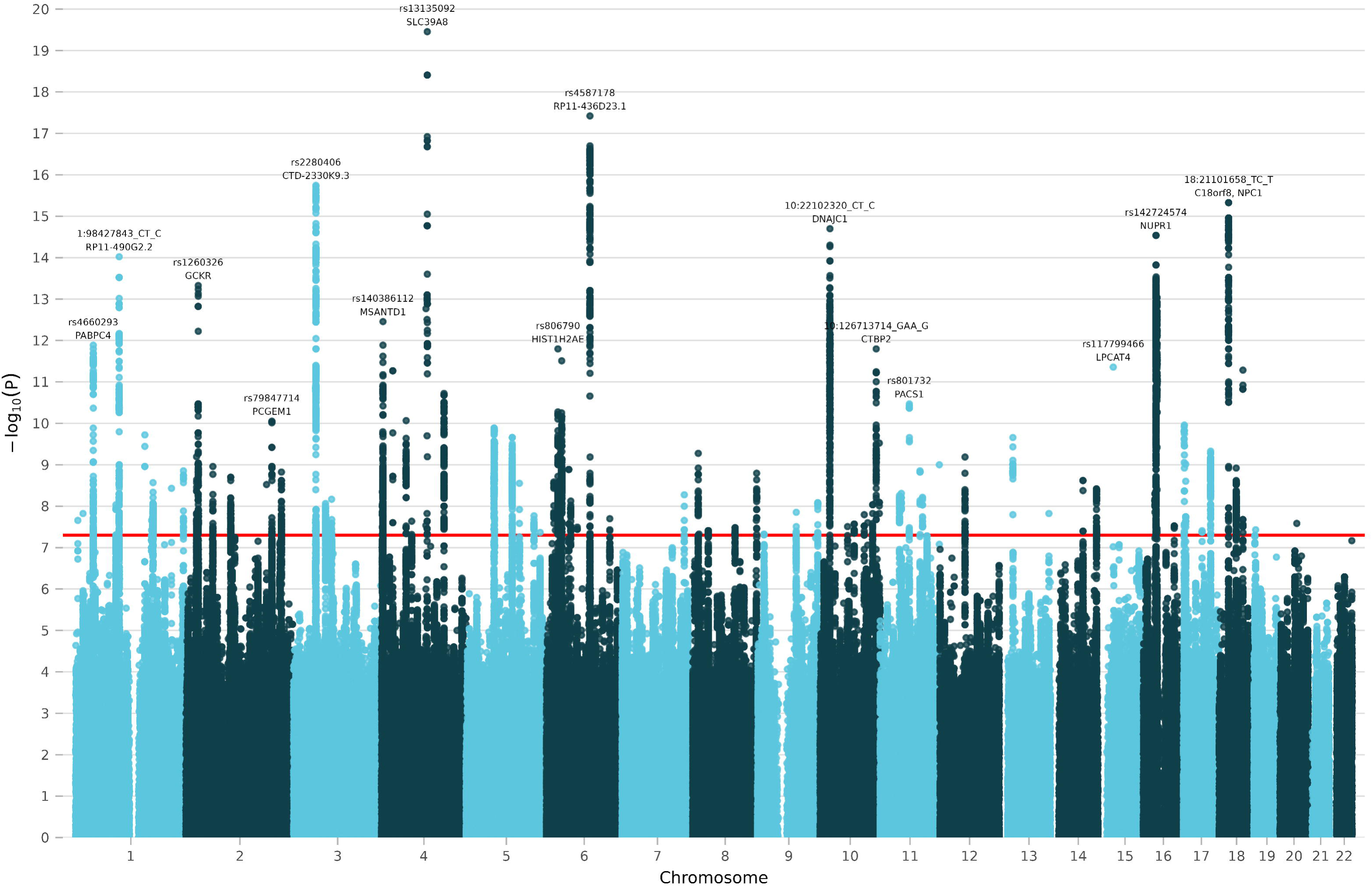
Manhattan plot for the polyexposure score for type 2 diabetes. Results of genome-wide association study performed on the polyexposure score for type 2 diabetes. Of the 19,400,443 SNPs plotted, 6,306 exceed the Bonferroni significance threshold (p < 5 x 10^-8^). The genomic inflation factor (λ_GC_) is 1.22.. Significant loci with p < 10^-10^ that were distant enough from other loci are labeled with their lead SNP and closest gene.

We also found a missense variant, rs1260326, strongly associated with higher PXS-T2D scores (beta = 0.01, risk allele: T). This locus maps to *GCKR* (Glucokinase regulatory protein, Table S3, Figure 2), expressed both in the liver and pancreatic beta cells. The variant has strong prior association in GWASs for T2D (OR = 1.1, p < 7 x 10^-52^) and concordant in direction with respect to the risk allele (e.g., higher odds for T2D for individuals with the T allele). Interestingly, the variant is also associated with clinical risk factors, such as total cholesterol (p < 5 x 10^-324^), triglycerides (p < 5 x 10^-324^) and fasting glucose (adjusted for BMI, p < 7 x 10^-91^). While the risk allele is associated with higher glucose levels (concordant with PXS-T2D and T2D), it is also associated with lower cholesterol and triglyceride levels.

Other top loci included rs4587178 (beta = 0.0017, p = 3.8 x 10^-18^), which was previously found to be associated with the first principal component of dietary patterns in the UK Biobank^23^ (p = 9.7 x 10^-10^) and is associated with higher urinary sodium excretion (p = 6.8 x 10^-9^) in the T2DKP. Interestingly, it was not associated with T2D itself (p = 0.09, OR = 1.008). The next top locus was rs2280406 (beta = -0.0136, p = 1.8 x 10^-16^), which has been associated with HDL cholesterol (p = 7.4 x 10^-56^), BMI (p = 5.90 x 10^-42^), and plasma C-reactive protein (2.25 x 10^-26^), but narrowly misses genome-wide significance for type 2 diabetes (p = 6.78 x 10^-8^). The locus maps to multiple genes, including *MST1R*, *CTD-2330K9.3*, *MON1A*, *TRAIP*, and *CAMKV*. 18:21101658_TC_T (p = 4.70 x 10^-16^) is also strongly associated with lower triglyceride levels (p = 8.34 x 10^-13^). Across the 342 genes associated with PXS-T2D, 42 (12.4%) had a previous association with T2D, as currently documented by the T2DKP.

### Body Mass Index, FTO locus, and PXS-T2D

A locus associated with *FTO*, rs9937521, known for its strong association with both obesity^24^ and T2D (p = 3.5 x 10^-80^ in the T2DKP), was significant in 8 of our component behavior GWAS including “White bread consumption”, “No recent dietary change”, and “Sleep Duration”.

However, none of the SNPs that mapped onto the *FTO* gene were identified as genome-wide significant in our PXS-T2D GWAS (most significant SNP: rs9937521, p = 4.3 x 10^-6^), although the SNP effect sizes in the *FTO* region were directionally consistent with their effects in T2D. SNPs found near the *FTO* region were significantly enriched for association with PXS-T2D (p = 3.9 x 10^-34^, Figure S3), with 19 SNPs reaching the threshold of suggestive significance (p < 10^-5^). Colocalization analyses of GWAS results in the *FTO* region show strong posterior probability that PXS-T2D has the same causal variant as the other behaviors for which *FTO* was found to be significant in (Figure S4). A full list of all genomic loci and genes associated with PXS-T2D is shown in Table S3.

BMI was by far the most replicated trait among our behaviors’ associated SNPs, followed by insomnia and T2D. Since the GWAS Catalog does not have standardized names for traits, we grouped the top 150 traits by replicated SNPs into 9 groups (traits within each group listed in Table S4). The most replicated trait group was “metabolic traits”, followed by “anthropometric traits” (mostly consisting of BMI and related metrics) and “cognitive/behavior traits” (see Tables S5 and S6).

### PXS-T2D exhibits genetic correlation with clinical risk factors and metabolic traits

We estimated the genetic correlation between the PXS-T2D and six CRFs (BMI, glucose, triglycerides, systolic blood pressure, HDL, and HbA1c; Figure 3). All genetic correlations were significant. PXS-T2D had a moderate mean absolute genetic correlation (mean |r_g_| = 0.25) across the six CRFs, with BMI being its strongest correlation (r_g_ = 0.57). We also estimated the same genetic correlations with our 25 component behaviors. “Usual walking pace” had the highest mean absolute genetic correlation (mean |r_g_| = 0.21) while also being the third most significant component of the PXS-T2D. For BMI, triglycerides, and HDL, the genetic correlation with the highest absolute value was with PXS-T2D (and not an individual behavior component).

**Fig. 3.**
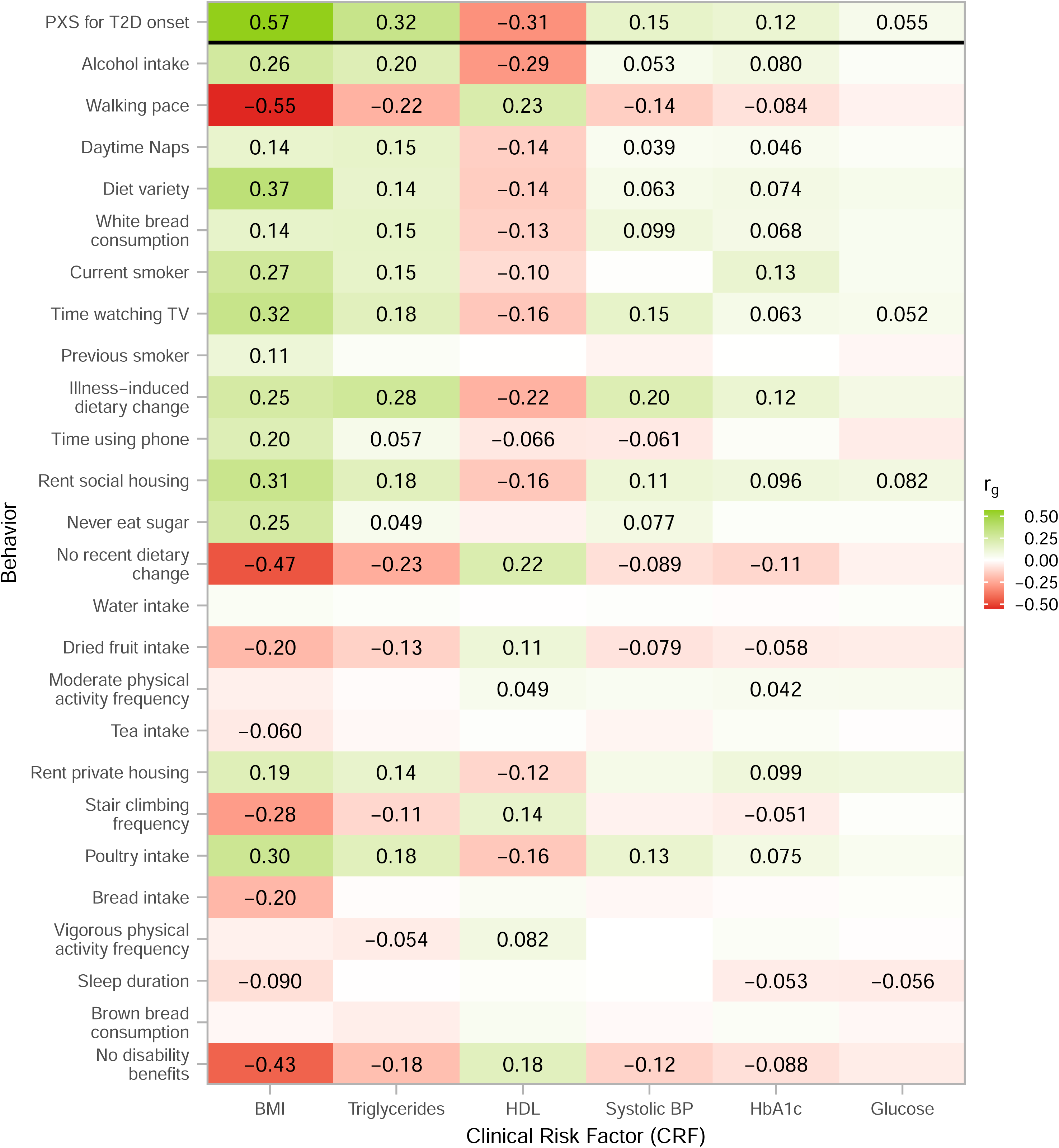
Genetic correlation between PXS-T2D plus individual behaviors and T2D clinical risk factors. Genetic correlations (r_g_) between T2D-associated behaviors (including PXS-T2D itself) and clinical risk factors for T2D as estimated by BOLT-REML. Only Bonferroni-adjusted significant genetic correlations have text displayed in the plot. Behaviors listed higher in the y-axis are also more strongly associated with incident T2D as reflected by their p-values in the PXS.

We also estimated the genetic correlation using LD-score correlation using PXS-T2D summary statistics and from summary statistics in the T2DKP (e.g., ^25^ Figure S8) to more broadly examine T2D-related physiological traits, including fasting insulin and proxies of insulin resistance (e.g., HOMA-IR). First, we found that LD-score-based genetic correlations between PXS-T2D and CRFs were not significantly different in the T2DKP vs. UKBB, with the exception of HbA1c (r_g_ = 0.123, [0.090, 0.156] in UK Biobank vs. r_g_ = 0.242, [0.192, 0.292] in T2DKP) and random glucose (r_g_ = 0.055, [0.0003, 0.1096] vs. r_g_ = 0.181, [0.1102, 0.253]). This is likely due to our inclusion criteria for the PXS generation: we removed individuals who had HbA1c values above 48 mmol/mol (see Methods). Furthermore, we found that the observational correlation between HbA1c and glucose is significantly smaller when individuals with higher baseline HbA1c are removed (r = 0.617, [0.615, 0.619] vs. r = 0.212, [0.209, 0.215]). We found that PXS-T2D and incident T2D were moderately genetically correlated (r_g_ = 0.474).

Next, we correlated the PXS-T2D with fasting glucose, fasting insulin, HbA1c, BMI, HOMA-IR, and HOMA-B. We found strong genetic correlations between PXS-T2D and HOMA-IR (r_g_ = 0.512, [0.360, 0.664]) and HOMA-B (r_g_ = 0.295, [0.181, 0.410]). On the other hand, we found weaker and non-significant correlations with fasting glucose (r_g_ = -0.006, [-0.091, 0.079]) and 2-hour glucose (r_g_ = 0.056, [-0.058, 0.170]). Compared to fasting glucose and 2-hour glucose, we found higher and significant correlations with fasting insulin (r_g_ = 0.143, [0.0599, 0.226]) and HbA1c (r_g_ = 0.168, [0.0949, 0.241]. Interestingly, BMI had a significantly stronger genetic correlation with PXS-T2D than with T2D itself (r_g_ = 0.57 vs. r_g_ = 0.42 respectively), while the opposite was true for glucose, HbA1c, HDL, and systolic blood pressure (Figure S5).

We provide a broad atlas of genetic correlation for traits in the T2DKP (Table S7, n = 628). In addition to BMI and HOMA-IR, the phenotypes with the largest genetic correlation with PXS-T2D included Non-Alcoholic Fatty Liver Disease ([NAFLD] r_g_ = 0.75, p = 1.4 x 10^-21^) and gastroesophageal reflux disease (r_g_ = 0.72, p < 1 x 10^-20^).

### Tissue expression of behavior-associated genes

Given that PXS-T2D shares genetic architecture with metabolic traits, we hypothesized that associated variant’s mapped genes should have similar expression patterns. After Bonferroni correction, the only tissue types of the 30 in GTEx to be significantly enriched in expression for any behavior’s or PXS-T2D’s associated genes are the brain and the pituitary (Figure 4, Figure S6), with the exception of the association between “Major dietary changes in the last 5 years because of illness” and testis tissue (p = 0.0183). Brain tissue was most strongly enriched for expression for the traits “Nap during day” (p = 5.10 x 10^-10^), “Sleep duration” (p = 4.11 x 10^-9^), and “Time spent watching television” (p = 6.89 x 10^-9^). Adjusting PXS-T2D for BMI still yielded significant expression enrichment in brain tissue (p = 6.42 x 10^-5^).

**Fig. 4.**
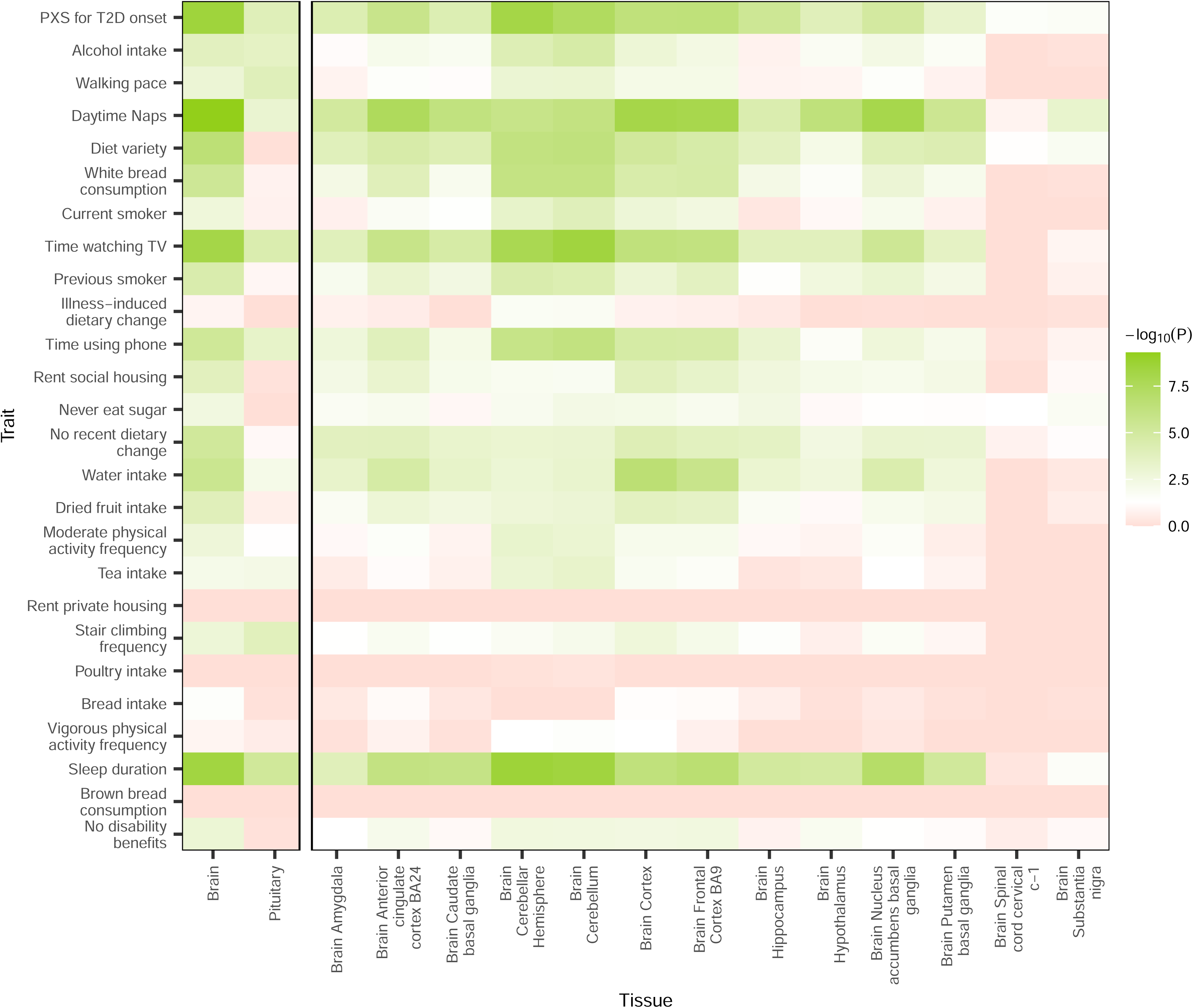
Enriched expression of behaviors’ associated genes across 30 tissue types. GWAS summary statistics for PXS-T2D and its 25 behaviors components were fed into FUMA to calculate the significance of differential expression in 30 tissue types and 53 specific tissues in GTEx. Within each of the two analyses, we further Bonferroni-adjusted the p-values for the number of traits tested. The heatmap colors were set such that p = 0.05 appeared as white, p > 0.05 appeared as red, and p < 0.05 appeared as green. Only tissue types (left section) and specific tissues (right section) with significant expression in at least one trait are shown and combined into one plot. The top row consists of tissue expressions for PXS-T2D, and behaviors listed higher in the y-axis are also more strongly associated with incident T2D as reflected by their p-values in the PXS. For the full expression heatmaps for all tissue types and specific tissues, see Figures S5 and S6 respectively.

We found similar results when looking across GTEx’s 53 specific tissues, some of which overlap with tissue types tested above. Only brain-related tissue, the pituitary, and the testis are enriched for expression in any behavior (Figure S7). Among brain tissues, PXS-T2D is most strongly enriched in the cerebellum (p = 3.97 x 10^-8^), frontal cortex (p = 4.39 x 10^-7^), and hippocampus (p = 4.21 x 10^-6^).

## Discussion

In summary, we interrogated the genetic determinants of cumulative exposure behaviors that are predictive of longitudinal type 2 diabetes. Findings from longitudinal diabetes studies have shown that risk may manifest 10 years or greater before onset^26^, but not readily apparent via glucose levels ^27^. Study of longitudinal exposures is important, but a challenge to understand how they exert their impact. Here, we conduct massive gene-by-exposure correlations to identify behavior exposures related to physiological components of T2D. Overall, we find (1) the genetic basis of a new latent behavior risk factor (the PXS-T2D) (2) the genetic architecture of the feature is connected with adiposity and insulin resistance, but not to glucose levels, complementary but distinct from current day T2D GWAS. We provide data resources for others to interrogate other components of T2D physiology.

T2D is caused by a combination of decreased insulin sensitivity (i.e., increase in insulin resistance) and secretion or a “decanalization” of the steady state.^28,29^ The underlying biological mechanisms by which behavioral exposure antecedents impact the steady state of insulin sensitivity and secretion, and ultimately T2D, are understudied. Re-analysis of GWAS of T2D and glycemic traits have recovered genetic signals that are associated with resistance and secretion to a varying degree.^11,30,31^

A major contributor to insulin resistance includes higher BMI.^32^ By comparing the genetic architecture of PXS-T2D with adiposity (e.g., BMI), cardiometabolic (e.g., triglycerides, cholesterol, blood pressure), and glycemic traits (e.g., hemoglobin A1c, glucose, insulin), we find that indeed the genetic architecture is largely shared between PXS-T2D and BMI (r_g_ = 0.57). Notably, the correlation between PXS-T2D and BMI is greater than the correlation between PXS-T2D and T2D itself (r_g_ = 0.47). We also found significant genetic correlation between PXS-T2D and HOMA-IR, HOMA-B, and triglyceride levels. In contrast, we observed smaller shared genetic architecture between glucose control and overall glucose levels. Our data on the PXS-T2D suggests that the role of PXS-captured behavior in T2D is via higher adiposity and potentially insulin resistance. Since these same clinical risk factors are associated with other detrimental health outcomes, estimation of genetic liability of cumulative unhealthy (or healthy) behavior can be potentially extended to diseases outside of diabetes.^33^

Human behavior, such as physical activity and dietary intake, is a result of the complex interactions between individuals agents and the specific environmental contexts they are in.^13,23,34,35^ Behavior is partly heritable,^36^ with estimates ranging from 5-15% in SNP-based heritability estimates^37^ to 30-60% in twin studies.^37–39^ We believe that the genetic variants associated with PXS-T2D are effective at capturing shared genetic architecture across behavioral traits. This builds on efforts that examine behavior factors one at a time in GWAS. For example, Merino and colleagues recently identified genetic variants associated with dietary intake behavior and cardiometabolic traits.^13^

Given that the PXS is a composite score of T2D risk-associated behavior, its genetic architecture could be understood as a measure of genetic liability of latent unhealthy behavior, or, in other words, behavior that is not directly measured by individual components, analogous “latent” traits in psychiatric research.^40,41^ For example, first, we found the GWAS-based heritability of the PXS-T2D to be 19%, greater than any one of the behavioral components alone. Second, exploring the genetic component of PXS-T2D offers insights that are not found when interrogating T2D directly. This is evidenced by PXS-T2D and T2D only sharing about half of their genetic architecture (r_g_ = 0.474) and that an overwhelming majority of genes associated with the former are not associated with latter phenotype.

We found functional enrichment of PXS-T2D in brain tissue, particularly in the cerebellum and cortex. This is inline with personality- and dietary-related variants being expressed in the brain,^13,42–46^ and recent studies have shown the important effect that brain-related variants have on metabolic health outcomes.^47^ The only other tissue type enriched in expression among PXS-T2D-associated genes was the pituitary gland.

Our paper comes with limitations. First, our GWAS had inflated genomic inflation factors (Table 1), which can be a sign of population stratification.^48^ We attempted to remedy this by using genetic principal components as covariates^49^ in linear mixed model software optimized for controlling for population structure,^50,51^ as well restricting our sample to self-reported “White British.” We therefore posit that the inflated lambdas in our GWAS are resulting from the high polygenicity^48^ often found in personality traits.^43,46^ A downside of filtering the genetic ancestral group used is that our results are less generalizable to other ancestral groups, compounded by the finding that lifestyle and psychological traits especially lack portability.^52^ In order to equitably share the benefits of research to all, we acknowledge that genomics research needs to diversify the samples used in studies.^53^ Furthermore, like all UK Biobank studies, our cohort is not entirely representative of the socioeconomic status and health outcomes of the general UK population.^54^ UK Biobank participants also lack extensive fasting or oral glucose tolerance testing measurements to estimate insulin sensitivity and secretion, two critical axes of diabetes etiology. In future studies, we aim to perform extensive exposome-wide assessment along the axes of fasting insulin and glucose to pin down what behaviors matter, and when they matter, for beta cell function and insulin resistance.

In this study, we attempt to disentangle the complex and correlated nature of behavioral exposures via a new computational construct, the polyexposure score (PXS). The PXS is an indicator of complex behavior exposures, summarizing activity, diet, and social exposure variables and is heritable, potentially impacting CRFs before T2D onset. Additionally, PXS may have implications for mental factors not previously linked to T2D, which merits further investigation. The trait correlations, associated genetic loci, and the tissues where variants impact expression largely align with those connected to adiposity. However, there are additional findings that include both established T2D risk factors (e.g., triglycerides) and potentially novel risk factors that could provide greater specificity and potency to existing models, such as complex associations in *GCKR*.^55,56^ Highlighting these novel genetic loci may offer new insights into the intricate relationship between PXS and T2D risk.

## Online Methods

All code and methods for reproducing the results of this paper is publicly available on our <u>github</u> repository. We deposit the PXS-T2D summary statistics in FigShare (doi: 10.6084/m9.figshare.24975564). The UK Biobank project number for this project was 2288; the Harvard University IRB approved of this study (IRB: IRB16-245).

### Study Participants

The UK Biobank (UKBB) is a cohort consisting of ∼500,000 individuals recruited across 22 assessment centers in the United Kingdom between 2006 and 2010.^57^ The UKBB assessed participants through questionnaires, physical measurements, and biological samples at the assessment center. Disease diagnoses were also attained via questionnaires and through linkages to participant health care provider data. Individuals were genotyped for over 800,000 single nucleotide polymorphisms (SNPs) using the UK BiLEVE Axiom Array or the UK Biobank Axiom Array. Genotypes were imputed to over 96 million SNPs using the Haplotype Reference Consortium, the UK10K panel, and the 1000 Genomes panel.^58^ In our analysis, we filtered the sample to the 455,813 individuals with self-identified ethnic background of “White British” in order to reduce population structure which can inflate genetic variant associations with a phenotype.^48^ For subsequent analyses, the remaining sample was randomly split into a training and validation group according to a 2:1 ratio.

### Prospective Type 2 Diabetes Definition

We defined incident T2D as whether an individual had an ICD10 code of E11.X (non-insulin-dependent diabetes; T2D) reported for the first time after their first assessment with the UKBB. Those with diagnosed diabetes before their first assessment or HbA1c values above 48 mmol/mol were removed from the analysis. For supplementary analyses, we also defined a broader binary T2D variable purely by E11.X reported or diabetes self-reported at any time point, acknowledging that this may include Type 1 Diabetes cases. We defined time to incident T2D by the number of days between the day of an individual’s first assessment and the day of their T2D diagnosis. A total of 388,452 participants remained, 14,294 of which developed T2D after the first assessment.

### Building a Polyexposure Risk Score (PXS) for T2D

We followed a similar procedure to that employed by He et al.^9^ to calculate the PXS for incident T2D on UK Biobank participants. Among He et al.’s 111 UKBB fields representing the exposome, e.g., physical exposures (e.g, air pollution), and “lifestyle” behavior variables (e.g., smoking behavior), with greater than 90% coverage in the UK Biobank, we selected 65 individual-level exposure behavior variables (variables that are measured on individuals as opposed to external measures often measured at the neighborhood or regional level), which are listed in Table S1. We only analyzed behavioral exposures ascertained in the first examination. We used PHESANT,^59^ a tool for standardizing UKBB diverse variable types (e.g., continuous, categorical, ordinal), to harmonize phenotypic data, which included dummy encoding categorical variables, yielding a total of 103 variables. Using a modified version of the software package PXStools,^10^ we performed an exposure-wide association study (ExWAS) to associate each of the 103 exposome variables with incident T2D using Cox Proportional Hazards regression, controlled for sex, age, and the first 40 genetic principal components (PCs) as covariates. Only exposure variables with an FDR-adjusted p-value below 0.05 and their related binarized variables were passed onto the next step (e.g., for categorical variables that were split into binary variables, only one of these variables needed to be significant for all others to be included). Next, we fit a multivariate regression model as a function of the 84 ExWAS identified fields (which mapped to a total of 98 variables) using LASSO until only independently significant (p < 0.05) variables remain, again also adjusting for age, sex, and the first 40 genetic PCs. The coefficient of each regression term allows us to compute the polyexposure score for type 2 diabetes (PXS-T2D) for individuals with complete data.

### Polygenic Risk Scores and Mendelian Randomization

In order to quantify the relationship between PXS-T2D and polygenic risk scores (PRS) for T2D, we extracted field 26285 from the UK Biobank, which contains a PRS for T2D for over 486,000 participants trained on external data by Thompson et al.^15^ We note that the PRS-T2D was not trained on the exact same definition of incident T2D that we used for our PXS. We then calculated the correlation between the PXS-T2D and PRS-T2D of an individual. Furthermore, we used the inverse-variance weighted (IVW) method in the MendelianRandomization R package^60^ to estimate causal relationships between PXS-T2D and incident T2D. We used significantly associated variants for each phenotype derived from our genome-wide association studies (described below) as instrumental variables for Mendelian randomization in each direction.

### Performing genome wide associations for PXS-T2D and behaviors

We used BOLT-LMM v2.3.2^51,61^ to perform a genome-wide association study (GWAS) on the PXS-T2D phenotype, the 25 behavioral phenotypes that composed it, and incident T2D itself. We filtered the full set of 96 million imputed SNPs to just those with a minor allele frequency (MAF) above 0.001 and an imputation information score (INFO) above 0.3, narrowing the set down to 19,400,443 markers. We used LD scores from the 1000 Genomes Project^62^ European cohort. Sex, age, the assessment center where data was collected, and the first 40 PCs were covariates. We calculated the lambda genomic inflation factor (λ_GC_) for each GWAS as the ratio of median ^2^ statistic to the expected median ^2^ statistic (∼0.455).

### Annotating GWAS variants to function

We used FUMA to annotate variants associated with PXS-T2D, which calls the MAGMA software and inputs GTEx data, which links genetic variants to tissue specific gene expression, and the GWAS Catalog, which is a database of documented variant-phenotype associations.^63–69^ We also extracted variant- and gene-level associations with the T2D phenotype on the Common Metabolic Disease Knowledge Portal^19^ (T2DKP, https://t2d.hugeamp.org/), which meta-analyzed 44 individual T2D GWAS. We also used the T2DKP to identify our significant variants’ associations with other biologically relevant phenotypes

### Estimating heritability of PXS-T2D and behavior variables

We used BOLT-REML for estimating narrow-sense GWAS-based heritability of the same set of PXS-T2D and 25 individual behavior constituent variables. We used the same SNP-filtering settings and covariates as in the BOLT-LMM GWAS. We used the --remlNoRefine flag in BOLT-REML, which speeds up computation time by a factor of 2 to 3 at the expense of standard errors being around 1.03 times greater.

### Computing genetic correlation between PXS-T2D and metabolic risk factors

We obtained clinical risk factors including Body Mass Index (BMI), non-fasting glucose, triglycerides, systolic blood pressure, and high density lipoprotein (HDL) cholesterol, and hemoglobin A1c (HbA1c). Genetic correlation is one tool to help elucidate causal relationships between phenotypes in lieu of an experiment or a randomized trial.^70^ Here, we wished to demonstrate the genetic correlation between PXS-T2D and T2D-related traits using observational association data and summary statistics.

Using individual-level data from participants of the UK Biobank, we used BOLT-REML to estimate the genetic correlation between pairs of phenotypes and T2D-PXS. We used identical SNP-filtering steps, covariates, and --remlNoRefine flag as in the BOLT-LMM GWAS analysis. We performed genetic correlations between T2D-PXS (and a BMI-adjusted PXS-T2D) and the six clinical risk factors listed above. We also used summary statistics data and pipelines from the T2DKP^20^ to estimate genetic correlation between PXS-T2D and fasting glucose, fasting insulin, 2-hour glucose, HOMeostatic Assessment of Insulin Resistance (HOMA-IR) and Beta-cell function (HOMA-B) phenotypes.^71^ HOMA-IR and -B are predicted estimates of insulin resistance and beta-cell function respectively.^72^ To estimate genetic correlation for summary statistics, we used LDsc^70,73^ with European LD scores as a reference.^62^ We also used the coloc R package to perform colocalization analysis to identify overlapping genetic signals between phenotypes in select regions^74^.

## Data Availability

All code and methods for reproducing the results of this paper is publicly available on our github repository. We deposit the PXS-T2D summary statistics in FigShare (doi: 10.6084/m9.figshare.24975564).

## Acknowledgements

We thank Jose Florez and Shamil Sunyaev for providing valuable feedback on the initial findings. This study was funded by NIEHS R01 ESS032470 and NIA RF1AG074372. We thank all participants and contributors of the UK Biobank for making this study possible.

## Author Contributions

Conceptualization, N.C and C.P.; Methodology, N.C and C.P.; Formal Analysis, N.C, P.S., and C.P.; Writing - Original Draft, N.C and C.P.; Writing - Review & Editing, N.C, Y.H., P.S., J.F., J.M.M., M.U., J.M, and C.P.; Visualization: N.C and C.P.; Supervision, C.P.; Project Administration, C.P.; Funding Acquisition, C.P.

## Figure Legends

**Fig. S1. Correlation matrix of the 25 behavioral exposures making up the PXS-T2D.** A heatmap matrix displaying the correlation between every pair of 25 behavior variables in the UK Biobank. The polycor R package was used to compute correlations between mixed data type variables. The data types of each variable are shown in Table S1. Correlation values are shown on the plot, with positive correlations shaded in green and negative correlations shaded in red. Lower magnitude correlations have a more muted color that is closer to white. The main diagonal is omitted due to all correlations being 1. The matrix is mirrored across the diagonal.

**Fig. S2. Correlation between PRS-T2D and PXS-T2D.** Scatterplot between polygenic risk score for T2D and polyexposure score for T2D for 264,704 individuals in the UK Biobank. Trend line for the two variables is shown. The standard PRS for T2D was used, meaning that the GWAS training data is external to the UK Biobank.

**Fig. S3. Enrichment of PXS-T2D SNP associations in the *FTO* region.** Manhattan plot for PXS-T2D associations within the *FTO* region (in dark green; additional 50 kb regions in each direction shown in light green). Despite zero SNPs exceeding the threshold for genome-wide significance (red line, p < 5 x 10^-8^), 19 SNPs surpass the suggestive significance threshold (blue line, p < 1 x 10^-5^). Horizontal segmented bars represent the mean -log_10_(P) for SNPs in the region. SNPs in the *FTO* region have a significantly higher mean -log_10_(P) than surrounding non-*FTO* SNPs (t = 13.5, p = 3.9 x 10^-34^).

**Fig. S4. Pairwise colocalization analysis of GWAS results in the *FTO* region across 25 behaviors and PXS-T2D.** Colocalization analysis of GWAS results in the *FTO* region (± 50 kb) among all pairs of 25 behaviors and PXS-T2D. The number in each tile is the posterior probability of the two traits having one common causal variant as implemented by the coloc.abf() function in the coloc R package. Axis labels that are bolded denote that the *FTO* gene was deemed to be significant for that trait, meaning that at least one SNP that maps onto the *FTO* gene had genome-wide significance. Note that some traits without such a SNP, like PXS-T2D, have a high probability of sharing a common variant with traits that are associated with *FTO*.

**Fig. S5. Comparison of genetic correlation between PXS-T2D and T2D.** Barplot comparison of the genetic correlation between clinical risk factors and PXS-T2D vs T2D. Error bars shown are 95% confidence intervals, and the asterisks denote non-overlap between intervals within each CRF. Data for T2D’s genetic correlations taken from the T2DKP. HDL is the only clinical risk factor with a negative genetic correlation with PXS-T2D and with T2D.

**Fig. S6: Enriched expression of behaviors’ associated genes across 30 tissue categories.** GWAS results for PXS-T2D and its 25 component behaviors were fed into FUMA to calculate the significance of differential expression in 30 tissue types in GTEx. We further Bonferroni-adjusted the p-values for the number of traits tested. The top row consists of tissue expressions for PXS-T2D, and behaviors listed higher in the y-axis are also more strongly associated with incident T2D as reflected by their p-values in the PXS.

**Fig. S7. Enriched expression of behaviors’ associated genes across 53 tissues.** GWAS results for PXS-T2D and its 25 component behaviors were fed into FUMA to calculate the significance of differential expression in 53 specific tissues in GTEx. Many specific tissues are also listed as tissue types in Figure S6. We further Bonferroni-adjusted the p-values for the number of traits tested. The top row consists of tissue expressions for PXS-T2D, and behaviors listed higher in the y-axis are also more strongly associated with incident T2D as reflected by their p-values in the PXS.

**Fig. S8. Genetic correlation between behaviors + PXS-T2D and select glycemic traits.** Genetic correlations (r_g_) between T2D-associated behaviors (including PXS-T2D itself) and select glycemic traits associated with T2D as estimated by LDsc. Only Bonferroni-adjusted significant genetic correlations have text displayed in the plot. Behaviors listed higher in the y-axis are also more strongly associated with incident T2D as reflected by their p-values in the PXS.

