## Supplementary Figures for "Assessing the genetic contribution of cumulative behavioral factors associated with longitudinal type 2 diabetes risk highlights adiposity and the brain-metabolic axis"

**Supplementary Tables**

**Table S1. UKB Field IDs and data types.**
**Table S2. Coefficient estimates (weights) of T2D-PXS model by behavior and covariate.**
**Table S3. T2D-PXS Genome-wide association study (GWAS) summary statistics.** CHR: chromosome number, BP: basepair in chromosome, A0: reference allele, A1: risk allele, MAF: minor allele frequency, beta: beta coefficient of GWAS, SE: standard error of beta, p: Pvalue, closest_gene: closest annotated genes to SNP.
**Table S4. GWAS catalog trait groupings.**
**Table S5. GWAS catalog trait count.**
**Table S6. GWAS catalog group count.**
**Table S7. Genetic correlation between PXS-T2D and 628 phenotypes using summary statistics from the T2DKP.** Phenotype Table: phenotype, SE: standard error of geentic correlation, P: p-value of genetic correlation, h2: GWAS-based heritability

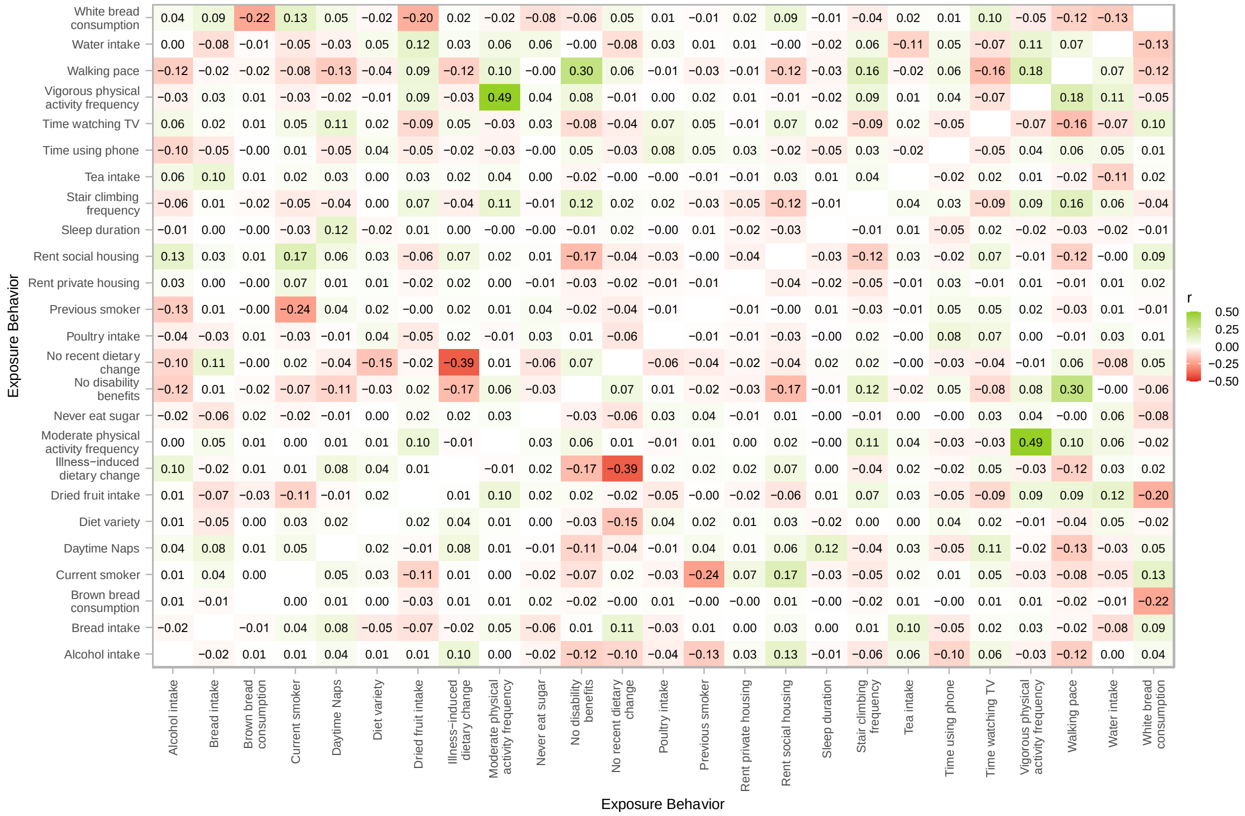


**Fig. S1. Correlation matrix of the 25 behavioral exposures making up the PXS-T2D.** A heatmap matrix displaying the correlation between every pair of 25 behavior variables in the UK Biobank. The polycor R package was used to compute correlations between mixed data type variables. The data types of each variable are shown in Table S1. Correlation values are shown on the plot, with positive correlations shaded in green and negative correlations shaded in red. Lower magnitude correlations have a more muted color that is closer to white. The main diagonal is omitted due to all correlations being 1. The matrix is mirrored across the diagonal.

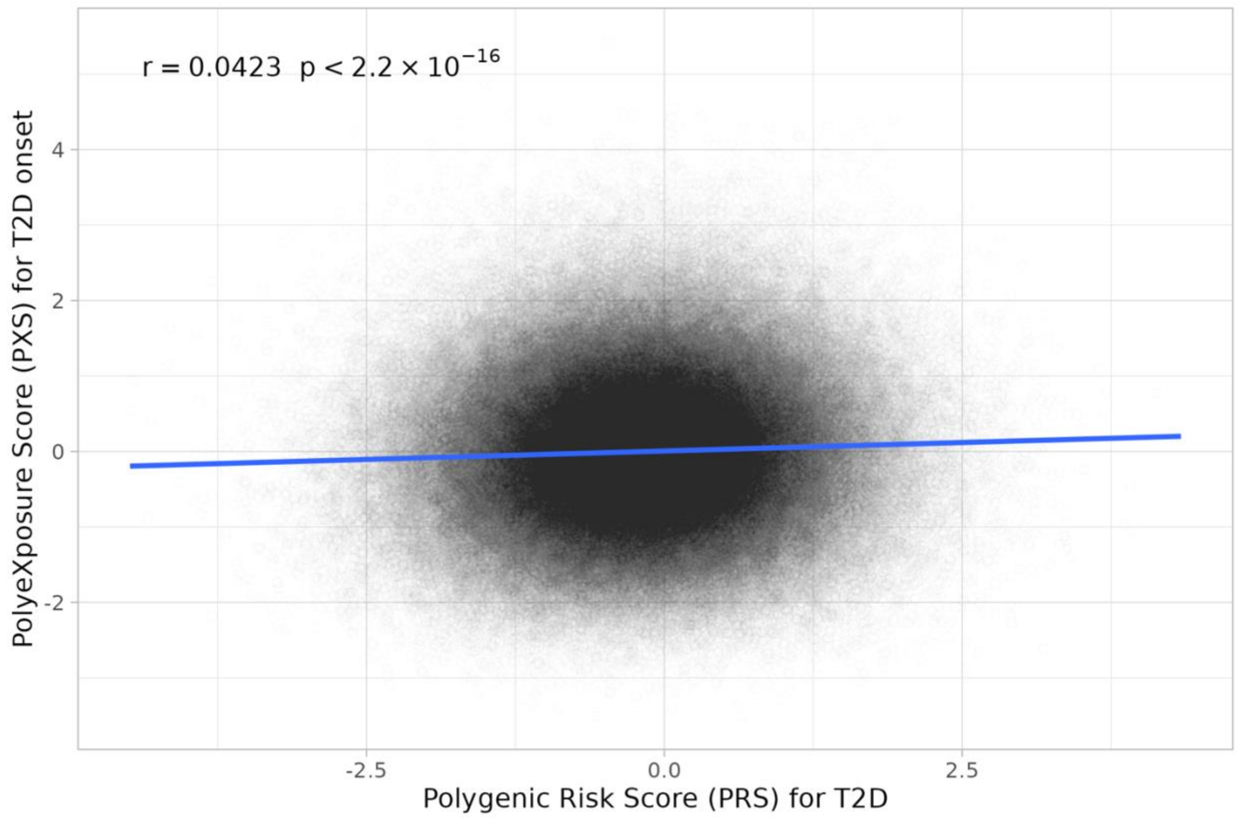


**Fig. S2. Correlation between PRS-T2D and PXS-T2D.** Scatterplot between polygenic risk score for T2D and polyexposure score for T2D for 264,704 individuals in the UK Biobank. Trend line for the two variables is shown. The standard PRS for T2D was used, meaning that the GWAS training data is external to the UK Biobank.

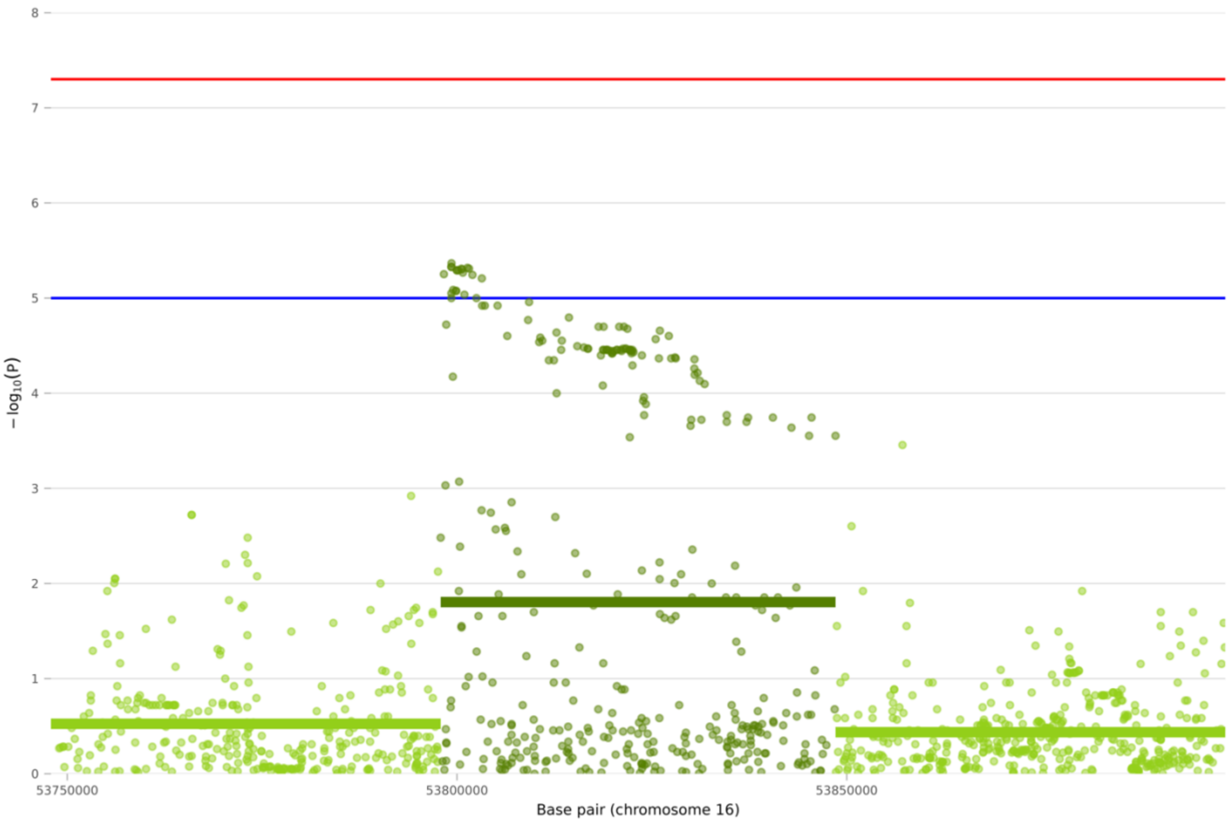


**Fig. S3. Enrichment of PXS-T2D SNP associations in the *FTO* region.** Manhattan plot for PXS-T2D associations within the *FTO* region (in dark green; additional 50 kb regions in each direction shown in light green). Despite zero SNPs exceeding the threshold for genome-wide significance (red line, $p < 5 \times 10^{-8}$), 19 SNPs surpass the suggestive significance threshold (blue line, $p < 1 \times 10^{-5}$). Horizontal segmented bars represent the mean $-\log_{10}(P)$ for SNPs in the region. SNPs in the *FTO* region have a significantly higher mean $-\log_{10}(P)$ than surrounding non-*FTO* SNPs ($t = 13.5$, $p = 3.9 \times 10^{-34}$).

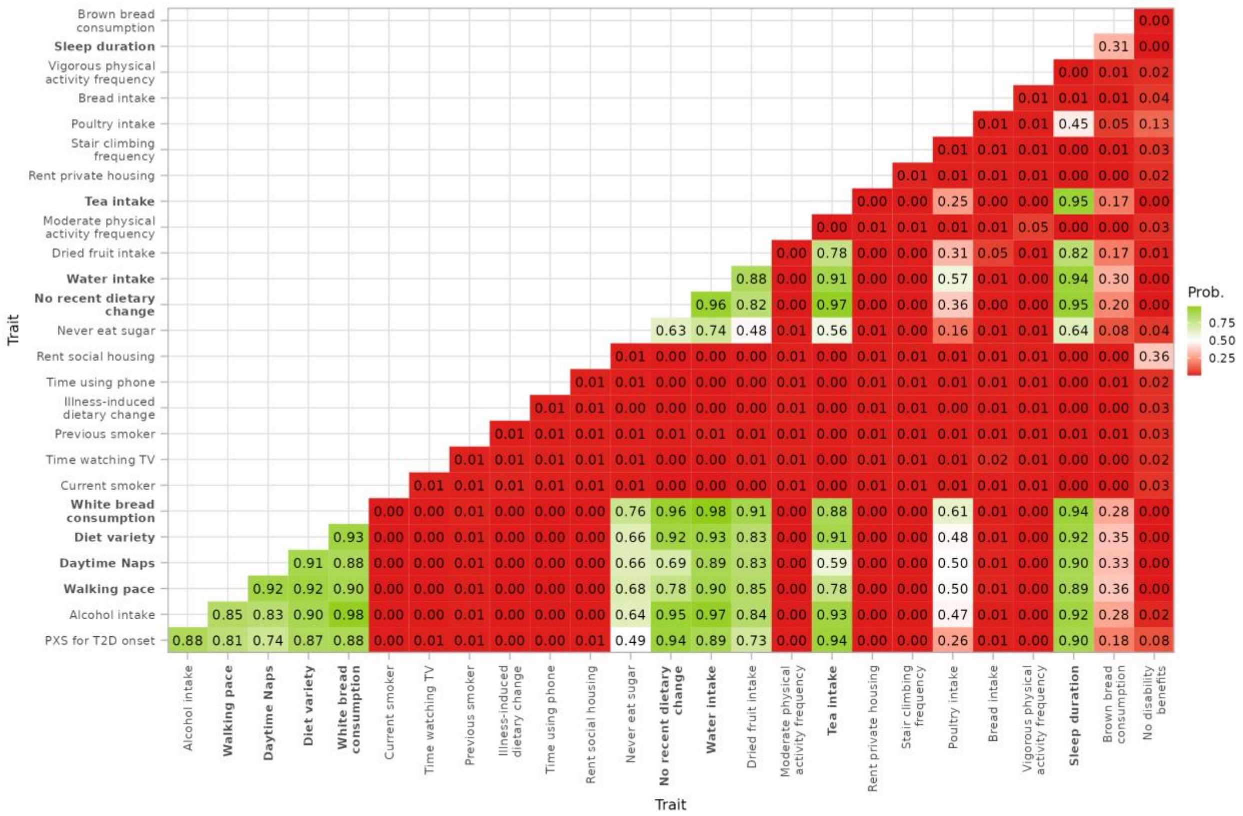


**Fig. S4. Pairwise colocalization analysis of GWAS results in the *FTO* region across 25 behaviors and PXS-T2D.** Colocalization analysis of GWAS results in the *FTO* region (± 50 kb) among all pairs of 25 behaviors and PXS-T2D. The number in each tile is the posterior probability of the two traits having one common causal variant as implemented by the coloc.abf() function in the coloc R package. Axis labels that are bolded denote that the *FTO* gene was deemed to be significant for that trait, meaning that at least one SNP that maps onto the *FTO* gene had genome-wide significance. Note that some traits without such a SNP, like PXS-T2D, have a high probability of sharing a common variant with traits that are associated with *FTO*.

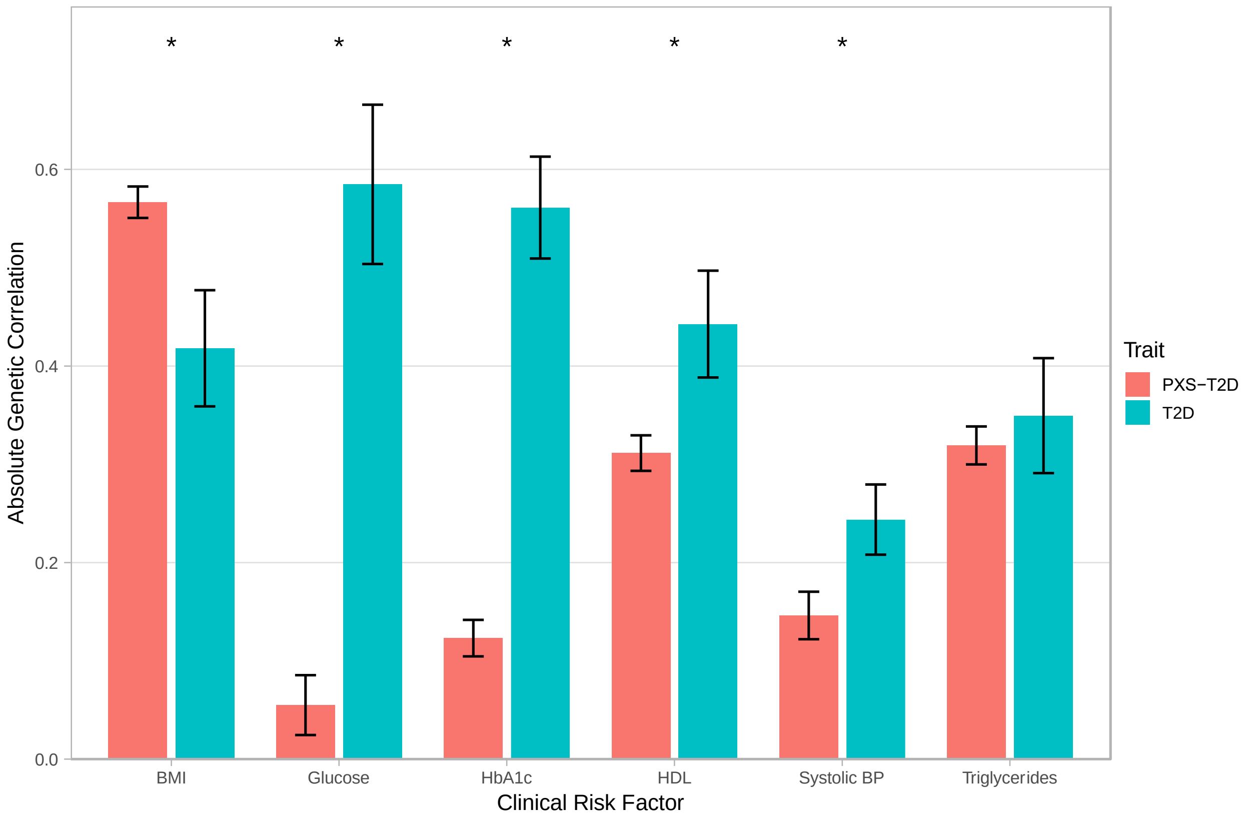


**Fig. S5. Comparison of genetic correlation between PXS-T2D and T2D.** Barplot comparison of the genetic correlation between clinical risk factors and PXS-T2D vs T2D. Error bars shown are 95% confidence intervals, and the asterisks denote non-overlap between intervals within each CRF. Data for T2D's genetic correlations taken from the T2DKP. HDL is the only clinical risk factor with a negative genetic correlation with PXS-T2D and with T2D.

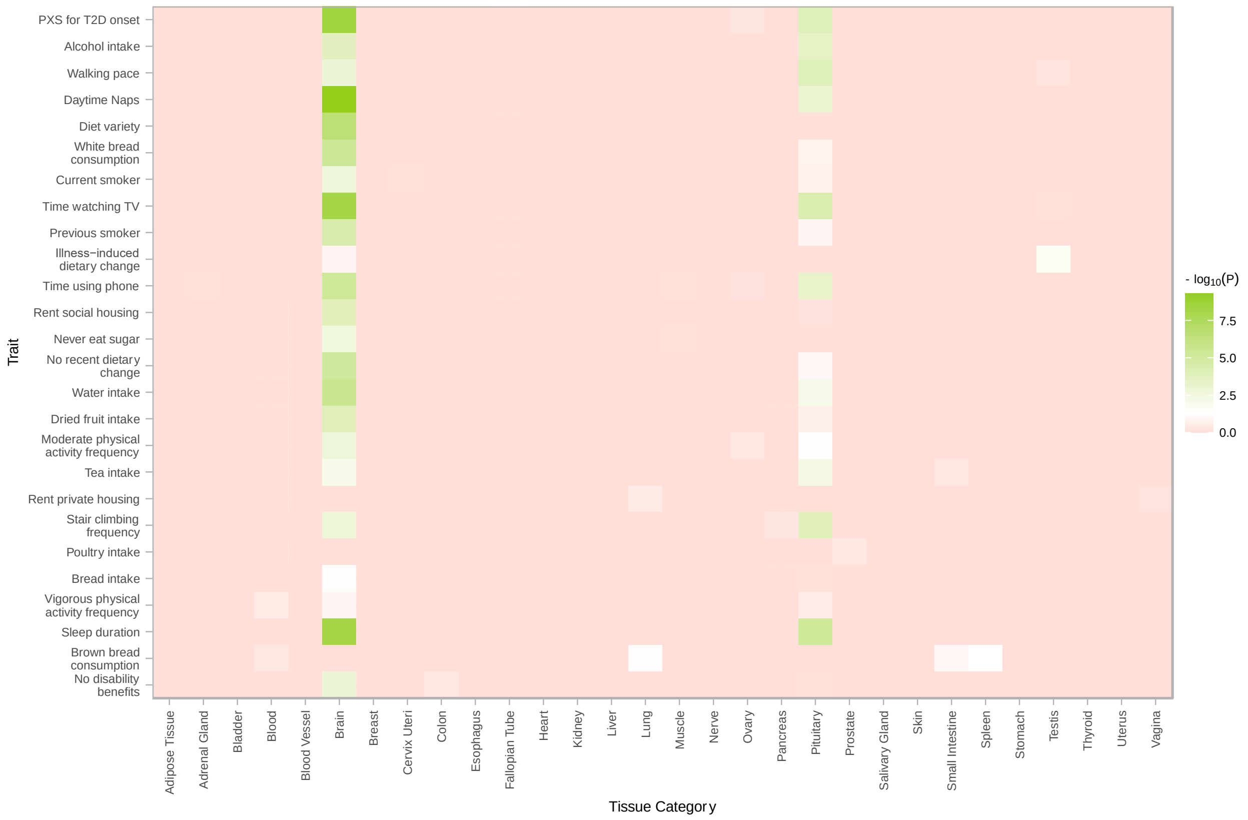


**Fig. S6: Enriched expression of behaviors' associated genes across 30 tissue categories.** GWAS results for PXS-T2D and its 25 component behaviors were fed into FUMA to calculate the significance of differential expression in 30 tissue types in GTEx. We further Bonferroni-adjusted the p-values for the number of traits tested. The top row consists of tissue expressions for PXS-T2D, and behaviors listed higher in the y-axis are also more strongly associated with incident T2D as reflected by their p-values in the PXS.

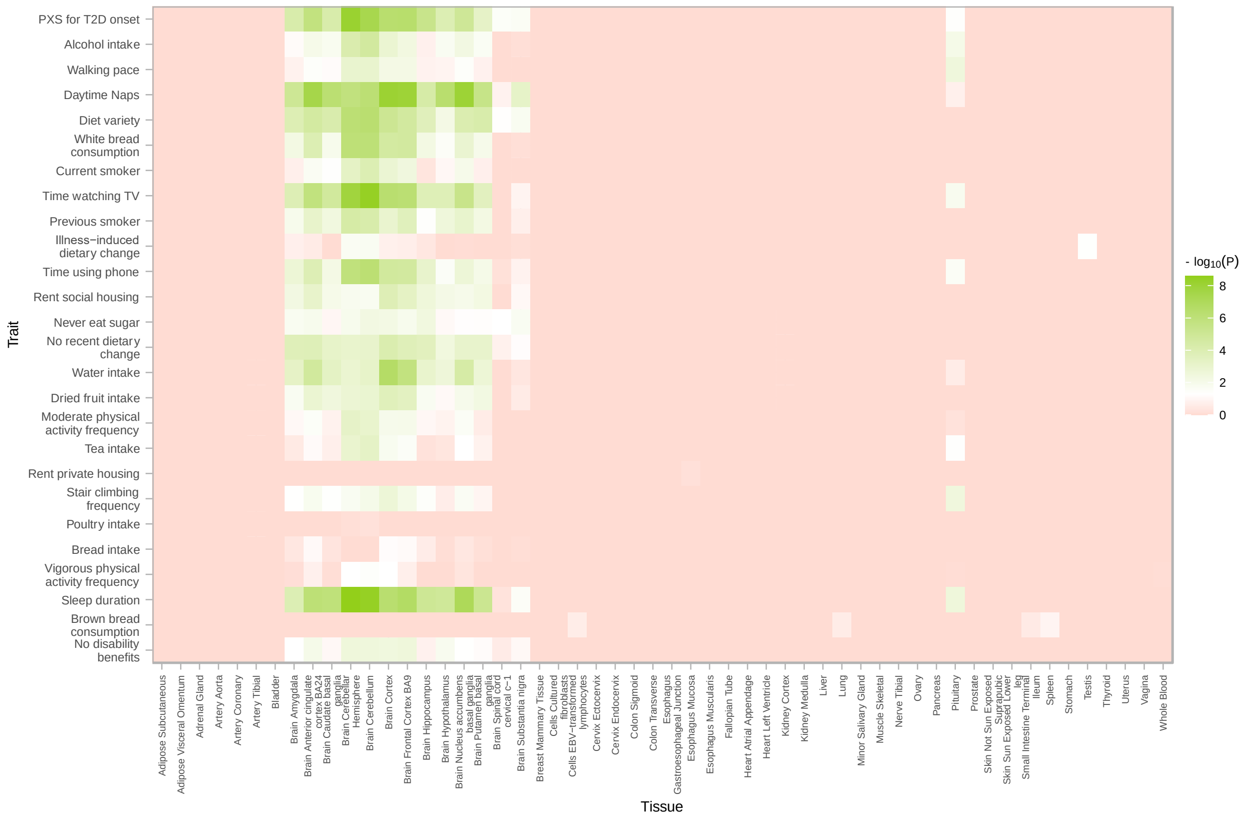


**Fig. S7. Enriched expression of behaviors' associated genes across 53 tissues.** GWAS results for PXS-T2D and its 25 component behaviors were fed into FUMA to calculate the significance of differential expression in 53 specific tissues in GTEx. Many specific tissues are also listed as tissue types in Figure S6. We further Bonferroni-adjusted the p-values for the number of traits tested. The top row consists of tissue expressions for PXS-T2D, and behaviors listed higher in the y-axis are also more strongly associated with incident T2D as reflected by their p-values in the PXS.

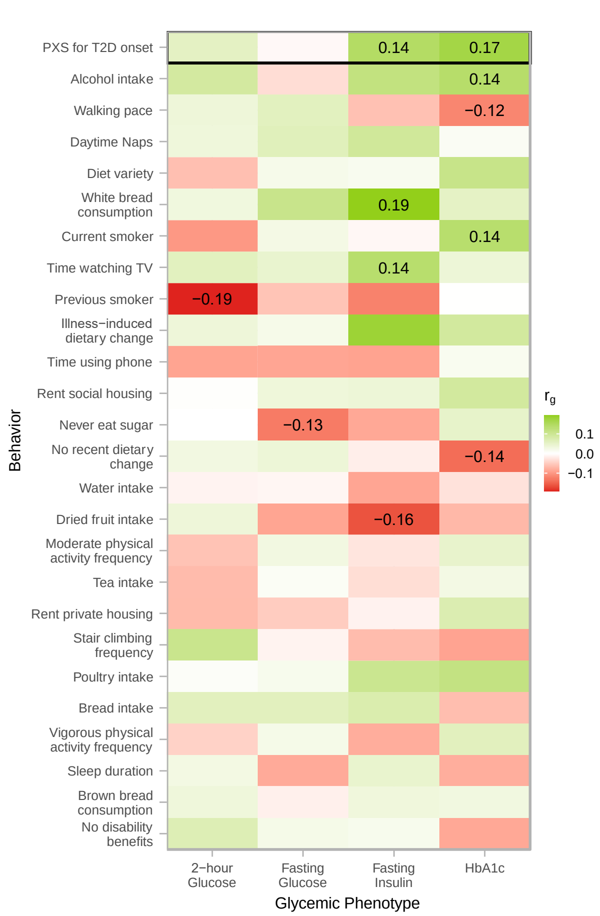


**Fig. S8. Genetic correlation between behaviors + PXS-T2D and select glycemic traits.** Genetic correlations ($r_g$) between T2D-associated behaviors (including PXS-T2D itself) and select glycemic traits associated with T2D as estimated by LDsc. Only Bonferroni-adjusted significant genetic correlations have text displayed in the plot. Behaviors listed higher in the y-axis are also more strongly associated with incident T2D as reflected by their p-values in the PXS.
